# Persistent Outpatient Oral Loop Diuretic Use and Clinical Outcomes According to Left Ventricular Volume Phenotype in Heart Failure With Preserved Ejection Fraction

**DOI:** 10.64898/2026.08.02.26359528

**Authors:** Dohyun Kim, Jiesuck Park, MinJung Bak, In-Chang Hwang, Hong-Mi Choi, Yeonyee E. Yoon, Goo-Yeong Cho

## Abstract

**Background:** Heart failure with preserved ejection fraction (HFpEF) is a heterogeneous syndrome, and patients with a small left ventricular (LV) volume phenotype exhibit greater preload dependency. Whether the association between persistent outpatient oral loop diuretic use and clinical outcomes differs by LV volume phenotype remains unclear.

**Methods:** We conducted a retrospective single-center cohort study of consecutive patients with heart failure and an LV ejection fraction ≥50% who underwent transthoracic echocardiography between 2009 and 2024. Patients were stratified by LV volume phenotype using sex-specific LV end-diastolic volume index cutoffs. Associations between outpatient oral loop diuretic use and HF hospitalization, worsening renal function, and all-cause death were evaluated using multivariable time-varying Cox proportional hazards models, including interaction testing by LV phenotype.

**Results:** Among 12,748 eligible patients, 1,437 had a small LV phenotype and 11,311 had a normal LV phenotype. Patients with a small LV phenotype initiated outpatient oral loop diuretics earlier and more frequently than those with a normal LV phenotype, while maintenance doses were comparable between groups. Outpatient oral loop diuretic use was associated with HF hospitalization in both phenotypes, with a significantly stronger association in the small LV group (hazard ratio [HR], 2.52; 95% confidence interval [CI], 1.68–3.78) than in the normal LV group (HR, 2.10; 95% CI, 1.77–2.49; P for interaction = 0.001). No significant interaction by LV phenotype was observed for worsening renal function or all-cause death. Fine–Gray competing-risk analyses showed a consistent interaction pattern.

**Conclusions:** Persistent outpatient oral loop diuretic use was more strongly associated with HF hospitalization in patients with a small LV phenotype than in those with a normal LV phenotype. LV volume phenotype may help identify patients who warrant closer monitoring and more individualized diuretic management.

**Clinical Perspective:** *What is new?:*

- In a large retrospective cohort of patients with heart failure with preserved ejection fraction, the association between persistent outpatient oral loop diuretic use and heart failure hospitalization differed according to left ventricular volume phenotype.
- This association was significantly stronger in patients with a small left ventricular volume than in those with a normal left ventricular volume, whereas left ventricular phenotype did not modify the associations with worsening renal function or all-cause death.

*What are the clinical implications?:*

- Patients with heart failure with preserved ejection fraction and a small left ventricular volume who require persistent outpatient oral loop diuretic therapy may warrant closer monitoring of volume status, symptoms, and treatment response.
- These observational findings should not discourage clinically indicated diuretic therapy for congestion but suggest that left ventricular volume assessment may help guide more individualized diuretic dosing and reassessment.

## INTRODUCTION

Heart failure (HF) with preserved ejection fraction (HFpEF) accounts for more than half of the overall HF population, and its prevalence continues to rise in parallel with population aging and the growing burden of cardiometabolic comorbidities.^1–3^ It is increasingly recognized as a heterogeneous syndrome rather than a single disease entity, and recent efforts have therefore focused on identifying distinct phenotypes with different pathophysiologic mechanisms, prognoses, and therapeutic responses.^4,5^ Among these, patients with a small left ventricular (LV) volume represent a distinct subgroup with unique hemodynamic features. Owing to a reduced LV cavity size and a leftward shift of the LV end-diastolic pressure–volume relationship, these patients operate on a relatively steep portion of the Frank–Starling curve and are highly preload-dependent, requiring adequate preload to maintain LV filling and stroke volume.^6,7^ Consequently, excessive preload reduction may result in a substantial decline in cardiac output and systemic perfusion.^8,9^

Loop diuretics are the mainstay therapy for symptom relief and volume control in HF and are widely prescribed in patients with HFpEF.^10,11^ Previous studies have reported variable and at times conflicting associations between chronic loop diuretic use and clinical outcomes.^12,13^ However, these analyses were largely conducted in unselected HFpEF populations and did not account for underlying differences in LV volume or preload dependency. This limitation may be particularly relevant in patients with a small LV volume, in whom sustained preload reduction could have greater hemodynamic consequences.^14,15^

Whether the association between persistent outpatient oral loop diuretic use and clinical outcomes differs according to LV volume phenotype remains unknown. In particular, it is unclear whether a small LV volume identifies a subgroup with greater susceptibility to persistent loop diuretic use. We therefore aimed to investigate the association between persistent outpatient oral loop diuretic use and clinical outcomes in patients with HFpEF and to determine whether this association differs according to LV volume phenotype.

## METHODS

### Study Population

This retrospective single-center cohort study included consecutive patients with HF who underwent transthoracic echocardiography (TTE) at Seoul National University Bundang Hospital between September 2009 and December 2024 (N=23,248). Patients were excluded if they had significant primary valvular heart disease (N = 1,681), left ventricular ejection fraction (LVEF) <50% (N = 5,128), HF hospitalization within 3 months before the index TTE (N = 1,122), follow-up duration <1 month (N = 863), unavailable LVEF or LV volume measurements (N = 186), or an outpatient oral loop diuretic prescription within 1 year before the index TTE (N = 1,520). Consequently, 12,748 patients comprised the final study cohort (**Figure 1**). The final cohort was stratified into small and normal LV volume groups according to LV end-diastolic volume index (LVEDVi), with small LV volume defined using the sex-specific lower limits of normal as an LVEDVi <34 mL/m² in men and <29 mL/m² in women.^16^ Of the final cohort, 1,437 patients were classified as having a small LV phenotype and 11,311 as having a normal LV phenotype. During follow-up, outpatient oral loop diuretic therapy was initiated in 238 patients in the small LV group and 1,331 patients in the normal LV group (**Figure 1**). The study protocol was approved by the Institutional Review Board of Seoul National University Bundang Hospital (IRB no. B-2605-1047-113), with a waiver of informed consent owing to the retrospective study design. D.K., J.P., and G.-Y.C. had full access to all study data and take responsibility for the integrity of the data and the accuracy of the data analysis.

**Figure 1.**
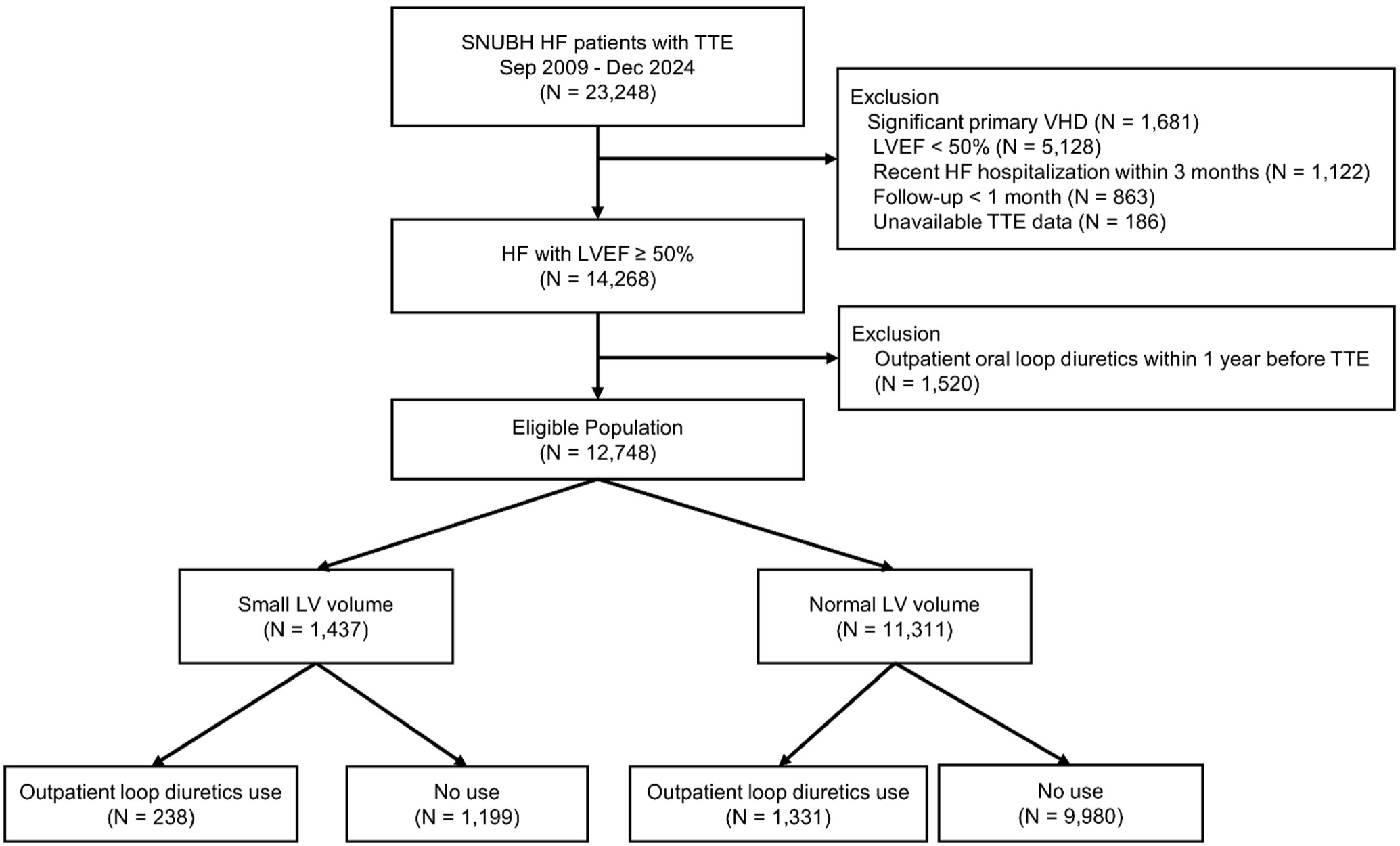
Study flow. Flow diagram of patient selection. Among consecutive patients with HF who underwent transthoracic echocardiography between September 2009 and December 2024, patients were sequentially excluded according to the predefined eligibility criteria. The final cohort was stratified into small and normal LV volume phenotypes according to left ventricular end-diastolic volume index and subsequently classified according to outpatient oral loop diuretic initiation during follow-up. HF, heart failure; LVEF, left ventricular ejection fraction; LV, left ventricle; LVEDVi, left ventricular end-diastolic volume index; SNUBH, Seoul National University Bundang Hospital; TTE, transthoracic echocardiography; VHD, valvular heart disease.

### Data Collection and Study Outcomes

Clinical data were retrospectively collected through a systematic review of the electronic medical records. Baseline variables included age, sex, body mass index, comorbidities (hypertension, diabetes mellitus, atrial fibrillation, prior myocardial infarction [MI], and chronic kidney disease [CKD]), serum creatinine, and NT-proBNP levels. Baseline use of renin–angiotensin system inhibitors (RASi), beta-blockers, mineralocorticoid receptor antagonists (MRAs), and sodium-glucose cotransporter-2 inhibitors (SGLT2i) was assessed within 3 months of the index echocardiographic examination.

All echocardiographic examinations were performed as part of routine clinical care by trained echocardiographers or cardiologists and interpreted by board-certified cardiologists specializing in echocardiography, in accordance with current guidelines.^16^ Echocardiographic variables included LVEF, LVEDVi, left ventricular mass index (LVMI), left atrial volume index (LAVI), and the ratio of early mitral inflow velocity to early diastolic mitral annular velocity (E/e′). LVEDVi was calculated as the LV end-diastolic volume indexed to body surface area.

Outpatient oral loop diuretic prescription records, including treatment initiation, discontinuation, and daily dose, were reviewed throughout follow-up. The doses of loop diuretics were standardized using the following equivalence: furosemide 40 mg and torasemide 20 mg were considered equivalent doses.^17^

The primary outcome was HF hospitalization. Secondary outcomes were worsening renal function and all-cause death. HF hospitalization was defined as admission with a primary diagnosis of HF accompanied by in-hospital intravenous loop diuretic use. Worsening renal function was defined as an increase in serum creatinine of ≥0.3 mg/dL from baseline that was sustained for at least 30 days. Clinical outcomes were ascertained from the index TTE until the occurrence of an outcome event, last follow-up, or death, as applicable.

### Statistical Analysis

The outcomes and the analytic approach, including stratification by LV phenotype, were prespecified prior to data analysis. Continuous variables are presented as medians with interquartile ranges and were compared using the Mann–Whitney U test. Categorical variables are presented as numbers with percentages and were compared using the chi-square test, as appropriate. Baseline characteristics were compared according to LV phenotype and, within each phenotype, according to subsequent outpatient oral loop diuretic use.

The cumulative incidence of outpatient oral loop diuretic initiation was estimated according to LV phenotype and compared at annual intervals throughout follow-up. Among patients receiving outpatient oral loop diuretics, dosing patterns were evaluated using prescription records, including mean daily dose, maximal daily dose, cumulative dose, and the number of prescriptions.

Associations between outpatient oral loop diuretic use and HF hospitalization, worsening renal function, and all-cause death were evaluated using multivariable time-varying Cox proportional hazards models. In these analyses, outpatient oral loop diuretic exposure was treated as a time-varying variable and updated from the date of treatment initiation. Hazard ratios (HRs) and 95% confidence intervals (CIs) were estimated. For the primary analysis, multivariable adjustment was performed for prespecified covariates, including age, sex, atrial fibrillation, diabetes mellitus, hypertension, CKD, prior MI, serum creatinine, LVMI, LAVI, E/e′, beta-blockers, MRAs, RASi, SGLT2i, and LVEF. LVEDVi was excluded from the multivariable models because it was used to define the small LV phenotype, whereas NT-proBNP was not included in the primary models because measurements were not available for all patients. To determine whether the associations between outpatient oral loop diuretic use and clinical outcomes differed according to LV phenotype, interaction terms between loop diuretic use and LV phenotype were included in the models.

Supportive Fine–Gray competing-risk analyses were performed for HF hospitalization and worsening renal function, with death treated as a competing event. Subdistribution hazard ratios (sHRs) with 95% confidence intervals (CIs) were estimated. As a sensitivity analysis, the primary multivariable models were repeated with additional adjustment for log-transformed NT-proBNP among patients with available measurements.

As an additional sensitivity analysis, the association between outpatient oral loop diuretic use and clinical outcomes was examined using an alternative exposure definition. The reference group comprised patients who never received outpatient oral loop diuretics or discontinued therapy within 45 days of initiation, whereas patients who continued therapy for ≥45 days were classified as persistent users. Under this definition, exposure status in the time-varying Cox models was updated from unexposed to exposed 45 days after treatment initiation. The same multivariable time-varying Cox and supportive Fine–Gray analyses were repeated using this exposure definition and the prespecified covariates.

All analyses were performed using Python (version 3.14). Two-sided P<0.05 was considered statistically significant.

## RESULTS

### Baseline Characteristics

Baseline characteristics according to LV phenotype are summarized in **Table 1**. Compared with patients with a normal LV volume, those with a small LV volume were older and more frequently male and had a higher prevalence of atrial fibrillation, diabetes mellitus, and chronic kidney disease, together with higher NT-proBNP levels. On echocardiography, patients with a small LV volume had lower LVMI and LAVI, whereas E/e′ was comparable between the groups.

**Table 1.** Baseline characteristics.

| Characteristic | Small LV<br>(n=1437) | Normal LV<br>(n=11311) | P value |
| --- | --- | --- | --- |
| *Demographics* |  |  |  |
| Age, years | 74.4 (64.7–81.3) | 68.2 (57.2–77.1) | <0.001 |
| Male | 785 (54.6) | 5274 (46.6) | <0.001 |
| BMI, kg/m<sup>2</sup> | 25.3 (22.9–27.7) | 24.8 (22.7–27.2) | <0.001 |
| *Clinical factors* |  |  |  |
| Hypertension | 938 (65.3) | 7506 (66.4) | 0.430 |
| Diabetes mellitus | 276 (19.2) | 1680 (14.9) | <0.001 |
| Atrial fibrillation | 295 (20.5) | 1332 (11.8) | <0.001 |
| Prior MI | 48 (3.3) | 346 (3.1) | 0.617 |
| CKD | 66 (4.6) | 388 (3.4) | 0.030 |
| *Laboratory* |  |  |  |
| Creatinine, mg/dL | 0.90 (0.75–1.07) | 0.81 (0.67–0.98) | <0.001 |
| NT-proBNP, pg/mL | 373 (87–1795) | 211 (66–1084) | <0.001 |
| *Echocardiography* |  |  |  |
| LVEF, % | 63.6 (59.9–67.3) | 62.9 (58.9–66.7) | <0.001 |
| LVEDVi, mL/m<sup>2</sup> | 28.0 (25.3–30.9) | 44.1 (38.2–51.9) | <0.001 |
| LVMI, g/m<sup>2</sup> | 82.0 (70.8–95.6) | 92.4 (78.8–108.1) | <0.001 |
| LAVI, mL/m<sup>2</sup> | 29.2 (23.5–38.8) | 34.0 (28.2–42.3) | <0.001 |
| E/e' | 9.8 (7.8–12.6) | 10.0 (8.0–12.7) | 0.360 |
| *Medications* |  |  |  |
| RASi | 209 (14.5) | 1873 (16.6) | 0.056 |
| Beta-blocker | 345 (24.0) | 2712 (24.0) | 1.000 |
| MRA | 100 (7.0) | 593 (5.2) | 0.008 |
| SGLT2i | 24 (1.7) | 231 (2.0) | 0.396 |

Data are presented as the median (interquartile range) for continuous variables and number (percentage) for categorical variables.
Abbreviations: BMI, body mass index; CKD, chronic kidney disease; E/e', ratio of early mitral inflow velocity to early diastolic mitral annular velocity; LAVI, left atrial volume index; LVEDVi, left ventricular end-diastolic volume index; LVEF, left ventricular ejection fraction; LVMI, left ventricular mass index; MI, myocardial ischemia; MRA, mineralocorticoid receptor antagonist; RASi, renin–angiotensin system inhibitor; SGLT2i, sodium-glucose cotransporter-2 inhibitor.

Within each LV phenotype, baseline characteristics were further compared according to subsequent outpatient oral loop diuretic use (**Table 2**). In both the small and normal LV volume groups, patients who subsequently received outpatient oral loop diuretics were older, more frequently had atrial fibrillation, and had higher NT-proBNP levels than non-users. They also had greater LVMI, LAVI, and E/e′ and were more frequently treated with beta-blockers and MRAs.

**Table 2.** Baseline characteristics by loop diuretic use.

| Characteristic | Small LV |  |  | Normal LV |  |  |
| --- | --- | --- | --- | --- | --- | --- |
|  | Loop Diuretics<br>(n=238) | No loop Diuretics<br>(n=1199) | P | Loop Diuretics<br>(n=1331) | No loop Diuretics<br>(n=9980) | P |
| ***Demographics*** |  |  |  |  |  |  |
| Age, years | 79.0 (73.4–83.1) | 73.2 (63.3–80.8) | <0.001 | 76.2 (70.0–82.2) | 66.8 (55.9–76.1) | <0.001 |
| Male | 116 (48.7) | 669 (55.8) | 0.054 | 570 (42.8) | 4704 (47.1) | 0.003 |
| BMI, kg/m<sup>2</sup> | 25.4 (22.8–28.1) | 25.2 (22.9–27.6) | 0.353 | 24.7 (22.4–27.3) | 24.8 (22.7–27.2) | 0.443 |
| ***Clinical factors*** |  |  |  |  |  |  |
| Hypertension | 113 (47.5) | 825 (68.8) | <0.001 | 649 (48.8) | 6857 (68.7) | <0.001 |
| Diabetes | 54 (22.7) | 222 (18.5) | 0.161 | 286 (21.5) | 1394 (14.0) | <0.001 |
| Atrial fibrillation | 91 (38.2) | 204 (17.0) | <0.001 | 364 (27.3) | 968 (9.7) | <0.001 |
| Prior MI | 8 (3.4) | 40 (3.3) | 1.000 | 68 (5.1) | 278 (2.8) | <0.001 |
| CKD | 15 (6.3) | 51 (4.3) | 0.226 | 99 (7.4) | 289 (2.9) | <0.001 |
| ***Laboratory*** |  |  |  |  |  |  |
| Creatinine, mg/dL | 0.96 (0.79–1.20) | 0.89 (0.74–1.04) | <0.001 | 0.90 (0.72–1.15) | 0.80 (0.66–0.97) | <0.001 |
| NT-proBNP,pg/mL | 980 (310–2522) | 236 (69–1314) | <0.001 | 933 (254–2673) | 143 (55–624) | <0.001 |
| ***Echocardiography*** |  |  |  |  |  |  |
| LVEF, % | 62.1 (57.7–66.0) | 63.8 (60.0–67.5) | <0.001 | 61.3 (56.9–65.9) | 63.1 (59.2–66.7) | <0.001 |
| LVEDVi, mL/m<sup>2</sup> | 27.2 (23.7–30.0) | 28.1 (25.5–31.1) | <0.001 | 44.1 (37.5–53.5) | 44.1 (38.3–51.7) | 0.417 |
| LVMI, g/m<sup>2</sup> | 85.0 (72.5–103.3) | 81.6 (70.0–94.6) | 0.009 | 103.2 (87.4–122.1) | 91.1 (78.0–106.2) | <0.001 |
| LAVI, mL/m<sup>2</sup> | 36.9 (28.9–51.3) | 28.5 (23.1–35.6) | <0.001 | 42.5 (32.9–58.3) | 33.3 (27.9–40.7) | <0.001 |
| E/e' | 11.4 (9.1–14.8) | 9.6 (7.7–12.3) | <0.001 | 12.3 (9.6–16.1) | 9.8 (7.8–12.2) | <0.001 |

| **Medications** |  |  |  |  |  |  |
| --- | --- | --- | --- | --- | --- | --- |
| RASi | 28 (11.8) | 181 (15.1) | 0.218 | 226 (17.0) | 1647 (16.5) | 0.689 |
| Beta-blocker | 86 (36.1) | 259 (21.6) | <0.001 | 435 (32.7) | 2277 (22.8) | <0.001 |
| MRA | 29 (12.2) | 71 (5.9) | <0.001 | 132 (9.9) | 461 (4.6) | <0.001 |
| SGLT2i | 4 (1.7) | 20 (1.7) | 1.000 | 20 (1.5) | 211 (2.1) | 0.168 |

Data are presented as the median (interquartile range) for continuous variables and number (percentage) for categorical variables.
Abbreviations: BMI, body mass index;; CKD, chronic kidney disease; ; E/e', ratio of early mitral inflow velocity to early diastolic mitral annular velocity; LAVI, left atrial volume index; LVEDVi, left ventricular end-diastolic volume index; LVEF, left ventricular ejection fraction; LVMI, left ventricular mass index; MI, myocardial ischemia; MRA, mineralocorticoid receptor antagonist; RASi, renin–angiotensin system inhibitor; SGLT2i, sodium-glucose cotransporter-2 inhibitor.

### Loop Diuretic Initiation and Dosing Patterns

Patients with a small LV phenotype initiated outpatient oral loop diuretics earlier and more frequently than those with a normal LV phenotype (**Figure 2** and **Table 3**). Within 1 year after the index echocardiogram, loop diuretics were initiated in 10.4% of patients with a small LV and 6.8% of those with a normal LV (P<0.001). The difference in initiation rates persisted throughout the 5-year follow-up period. Despite the earlier and more frequent initiation of loop diuretics in the small LV group, the mean maintenance daily dose remained comparable between the two groups throughout follow-up. Furthermore, among patients initiating therapy within 1 year, the mean daily dose, maximal daily dose, cumulative dose, and number of prescriptions were comparable between groups (**Table 3**).

**Figure 2.**
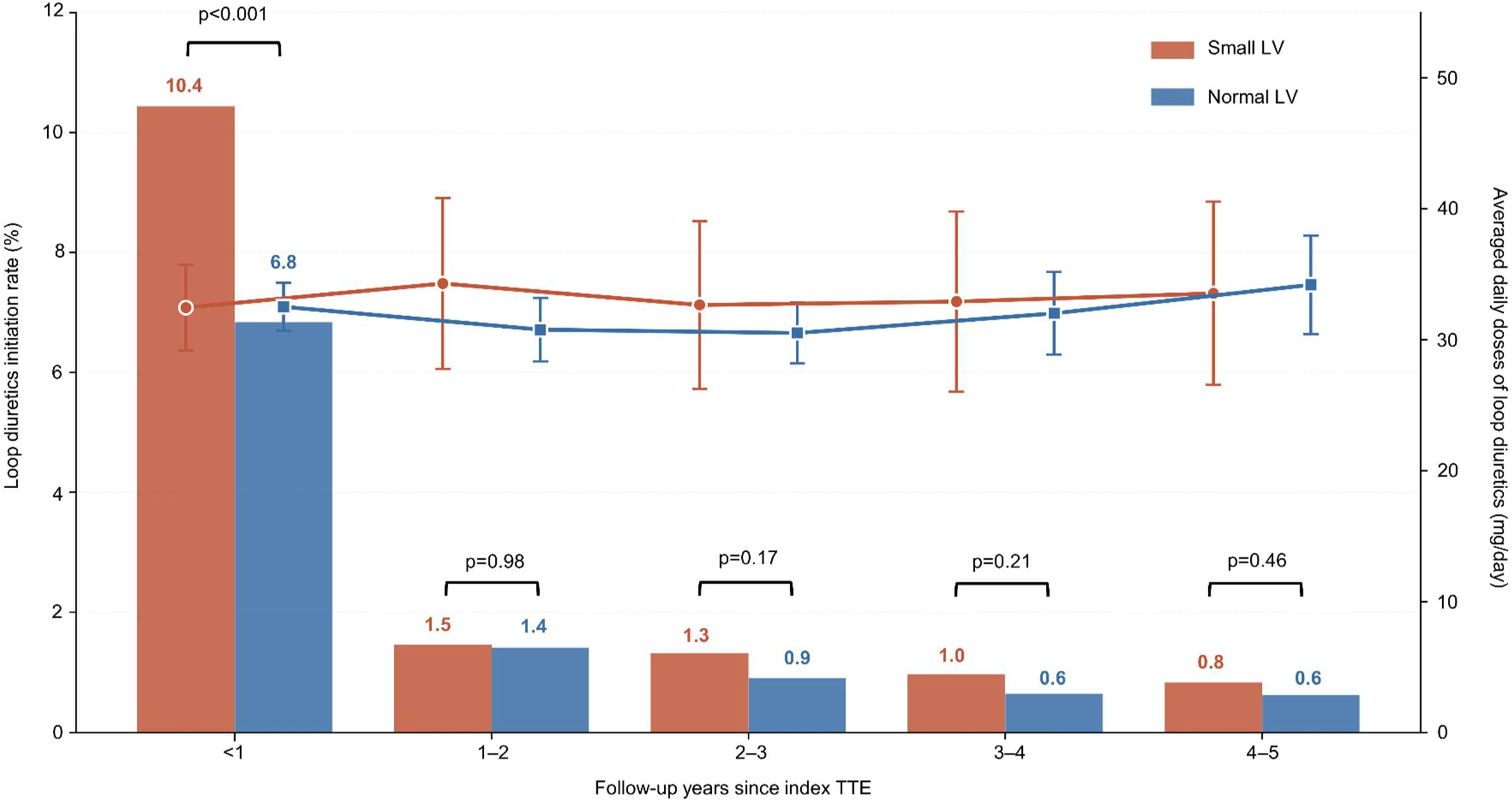
Initiation and dosing patterns of outpatient oral loop diuretics according to LV volume phenotype. Bars (left y-axis) represent the proportion of patients initiating outpatient oral loop diuretics during each follow-up interval according to LV phenotype. Lines (right y-axis) represent the mean daily loop diuretic dose, expressed as the furosemide-equivalent dose in mg/day, among treated patients during the corresponding follow-up intervals. LV, left ventricle; TTE, transthoracic echocardiography.

**Table 3.** Loop diuretic initiation and dosing patterns according to LV size.

| **Cumulative incidence of loop diuretic initiation, n (%)** |  |  |  |
| --- | --- | --- | --- |
| **Time since index echo** | **Small LV** | **Normal LV** | **P value** |
| Within 1 year | 150 (10.4) | 774 (6.8) | <0.001 |
| Within 2 years | 171 (11.9) | 934 (8.3) | <0.001 |
| Within 3 years | 190 (13.2) | 1037 (9.2) | <0.001 |
| Within 4 years | 204 (14.2) | 1110 (9.8) | <0.001 |
| Within 5 years | 216 (15.0) | 1181 (10.4) | <0.001 |
| **Loop diuretic dose\* within 1 year among users** |  |  |  |
| **Dose metric** | **Small LV** | **Normal LV** | **P value** |
| Mean daily dose, mg/day | 27 (20–40) | 24 (20–40) | 0.207 |
| Max daily dose, mg/day | 40 (20–40) | 40 (20–40) | 0.284 |
| 1-year cumulative dose, mg | 4,000 (1,120–8,680) | 3,950 (1,200–8,960) | 0.780 |
| No. of prescriptions | 3 (1–5) | 3 (1–5) | 0.962 |

\*Furosemide-equivalent doses were calculated as twice the torasemide dose.

### Clinical Outcomes

In multivariable time-varying Cox analyses, outpatient oral loop diuretic use was associated with a higher risk of HF hospitalization in both LV phenotypes (**Figure 3A**). The association was significantly stronger in patients with a small LV (HR, 2.52; 95% CI, 1.68–3.78) than in those with a normal LV (HR, 2.10; 95% CI, 1.77–2.49; P for interaction = 0.001). Consistent findings were observed in the Fine–Gray analysis treating death as a competing event, with a stronger association in the small LV group (sHR, 3.12; 95% CI, 2.38–4.10) than in the normal LV group (sHR, 1.53; 95% CI, 1.44–1.62; P for interaction <0.001). Loop diuretic use was also associated with worsening renal function in both LV phenotypes, but the association did not differ significantly according to LV phenotype in either the time-varying Cox model (small LV: HR, 1.85; 95% CI, 1.29–2.65; normal LV: HR, 2.68; 95% CI, 2.31–3.10; P for interaction = 0.627) or the Fine–Gray model (small LV: sHR, 1.74; 95% CI, 1.31–2.30; normal LV: sHR, 1.39; 95% CI, 1.30–1.49; P for interaction = 0.106) (**Figure 3B**). Similarly, loop diuretic use was associated with all-cause death in both LV phenotypes, without a significant interaction according to LV phenotype (small LV: HR, 1.74; 95% CI, 1.22–2.48; normal LV: HR, 2.11; 95% CI, 1.81–2.45; P for interaction = 0.056) (**Figure 3C**). Overall, LV phenotype significantly modified the association between outpatient oral loop diuretic use and HF hospitalization, but not the associations with worsening renal function or all-cause death.

**Figure 3.**
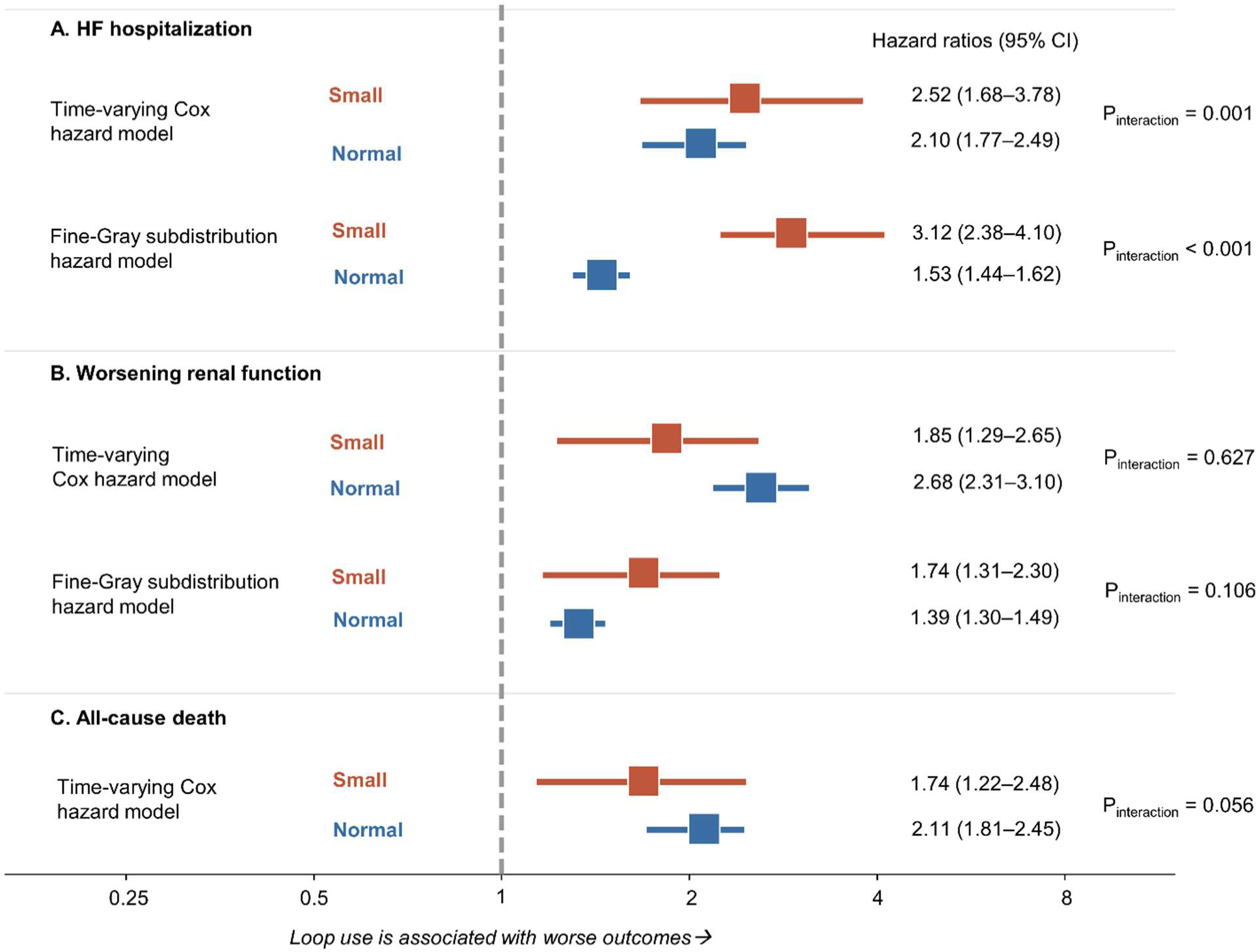
Associations between outpatient oral loop diuretic use and clinical outcomes according to LV volume phenotype. Forest plots showing hazard ratios with 95% confidence intervals for HF hospitalization (A), worsening renal function (B), and all-cause death (C) according to LV phenotype. HRs were estimated using multivariable time-varying Cox proportional hazards models. For HF hospitalization and worsening renal function, subdistribution hazard ratios from Fine–Gray competing-risk models with death treated as a competing event are additionally presented. P values for interaction indicate whether the association between outpatient oral loop diuretic use and each clinical outcome differed according to LV phenotype. CI, confidence interval; HF, heart failure; HR, hazard ratio; LV, left ventricle; sHR, subdistribution hazard ratio.

Additional adjustment for log-transformed NT-proBNP yielded findings consistent with the primary analysis (**Figure S1**). The association with HF hospitalization remained significantly stronger in patients with a small LV than in those with a normal LV in both the time-varying Cox model (HR, 2.24; 95% CI, 1.43–3.51 versus HR, 1.55; 95% CI, 1.28–1.89; P for interaction = 0.005) and the Fine–Gray model (sHR, 3.20; 95% CI, 2.27–4.50 versus sHR, 1.54; 95% CI, 1.40–1.69; P for interaction <0.001). No significant interaction according to LV phenotype was observed for worsening renal function or all-cause death.

The sensitivity analysis using persistent outpatient oral loop diuretic use for ≥45 days as the exposure definition also yielded findings consistent with the primary analysis (**Figure S2**). Persistent use remained more strongly associated with HF hospitalization in patients with a small LV than in those with a normal LV in both the time-varying Cox model (HR, 2.68; 95% CI, 1.63–4.42 versus HR, 2.24; 95% CI, 1.81–2.76; P for interaction = 0.003) and the Fine– Gray model (sHR, 2.93; 95% CI, 2.10–4.08 versus sHR, 1.69; 95% CI, 1.55–1.84; P for interaction <0.001). No significant interaction according to LV phenotype was observed for worsening renal function or all-cause death.

## DISCUSSION

In this retrospective cohort of patients with HFpEF, patients with a small LV phenotype initiated outpatient oral loop diuretics earlier and more frequently than those with a normal LV phenotype, despite receiving comparable maintenance doses. The association between persistent outpatient oral loop diuretic use and clinical outcomes differed according to LV phenotype. Although outpatient oral loop diuretic use was associated with HF hospitalization in both LV phenotypes, this association was significantly stronger in patients with a small LV phenotype. In contrast, LV phenotype did not modify the associations of loop diuretic use with worsening renal function or all-cause death. To the best of our knowledge, this is the first study to specifically evaluate the prognostic implications of persistent outpatient oral loop diuretic use according to LV volume phenotype in HFpEF.

The present findings are physiologically plausible given the unique hemodynamic characteristics of the small LV phenotype. Recent invasive hemodynamic studies have demonstrated that patients with HFpEF and smaller LV volumes exhibit increased LV diastolic stiffness together with a leftward shift of the end-diastolic pressure–volume relationship, resulting in reduced LV filling reserve.^18^ Consequently, these patients may be more vulnerable to reductions in preload. Given that maintenance loop diuretic doses were comparable between the LV phenotypes, the stronger association with HF hospitalization in the small LV group is unlikely to be explained solely by greater diuretic intensity. Rather, it may reflect greater susceptibility to the hemodynamic consequences of sustained preload reduction in patients with limited ventricular filling reserve.

Previous observational studies have consistently reported that chronic loop diuretic use or higher loop diuretic doses are associated with adverse outcomes in patients with chronic HF, including those with HFpEF.^12,13^ However, these studies primarily evaluated overall treatment effects and were unable to determine whether specific HF phenotypes exhibit differential susceptibility to loop diuretic therapy. In addition, interpretation of these observational findings has been challenged by confounding by indication, as patients receiving loop diuretics generally have more advanced disease and greater congestion severity. To address this limitation, our study sought to mitigate confounding by indication through prespecified stratification by LV phenotype, comprehensive multivariable adjustment, time-varying exposure modeling, and supportive Fine–Gray competing-risk analyses. The present findings extend the existing evidence by demonstrating that the association between persistent outpatient oral loop diuretic use and HF hospitalization differs according to LV volume phenotype. Furthermore, the observed interaction was specific to HF hospitalization and was not observed for worsening renal function or all-cause death, a pattern more consistent with phenotype-specific HF vulnerability than with generalized disease severity alone.

NT-proBNP was not included as a covariate in the primary multivariable model because measurements were unavailable in approximately 60% of patients. Sensitivity analyses with additional adjustment for log-transformed NT-proBNP among patients with available measurements yielded findings consistent with the primary analysis. A further sensitivity analysis distinguishing persistent use from no or transient use also reproduced the primary interaction pattern. Together, these analyses support the robustness of the observed association between persistent loop diuretic exposure and HF hospitalization according to LV phenotype.

The current study findings have potential clinical implications for the management of HFpEF. Current HF guidelines recommend loop diuretics as the cornerstone of symptomatic congestion relief regardless of HF phenotype.^10,11^ However, our findings suggest that patients with a small LV phenotype may represent a subgroup in whom sustained outpatient oral loop diuretic therapy warrants closer clinical monitoring. Rather than discouraging appropriate decongestion, these results support an individualized treatment strategy that incorporates LV volume phenotype when balancing relief of congestion against the potential consequences of excessive preload reduction. Accordingly, loop diuretics need not be avoided when clinically indicated for congestion, but persistent therapy may warrant more frequent reassessment of symptoms, volume status, and treatment response in patients with a small LV phenotype.

Several limitations should be acknowledged. First, this was a retrospective observational study in which outpatient oral loop diuretic therapy was prescribed according to clinical judgment rather than a standardized protocol. Therefore, residual confounding, including confounding by indication, cannot be completely excluded despite multivariable adjustment and time-varying exposure analyses. Second, prescription records were used to define loop diuretic exposure and may not capture actual medication adherence. Third, longitudinal data on congestion status, natriuretic peptide levels, and hemodynamic profiles were not systematically available, limiting more granular characterization of the mechanisms underlying the observed associations. Finally, this was a single-center study conducted in an East Asian population, which may limit the generalizability of the findings. Prospective studies are needed to validate these phenotype-specific associations and determine whether LV volume phenotype can help guide individualized outpatient diuretic management in patients with HFpEF.

In conclusion, patients with HFpEF and a small LV phenotype demonstrated a significantly stronger association between persistent outpatient oral loop diuretic use and HF hospitalization despite comparable maintenance loop diuretic doses. These findings suggest that LV volume phenotype may represent an important consideration when individualizing long-term outpatient diuretic therapy in patients with HFpEF.

## Author contributions

D.K. and J.P. contributed to conceptualization, methodology, formal analysis, investigation, data curation, and writing—original draft, and contributed equally as co-first authors. G.-Y.C. contributed to conceptualization, methodology, supervision, project administration, and writing—review and editing, and served as corresponding author. M.B., I.-C.H., H.-M.C., and Y.E.Y. contributed to conceptualization, and writing—review and editing. All authors have read and approved the final version of the manuscript

## Data availability

The data underlying this study cannot be made publicly available because of ethical restrictions imposed by the institutional review board, as public disclosure could compromise patient confidentiality and privacy. Requests for access to a minimal anonymized dataset may be directed to the corresponding author.

## Supplemental Material

Figure S1. Sensitivity analysis of the associations between outpatient oral loop diuretic use and clinical outcomes after additional adjustment for NT-proBNP, according to LV volume phenotype

Figure S2. Sensitivity analysis of the associations between persistent outpatient oral loop diuretic use and clinical outcomes according to LV volume phenotype

## Non-standard Abbreviations and Acronyms

CI: confidence interval
CKD: chronic kidney disease
E/e′: ratio of early mitral inflow velocity to early diastolic mitral annular velocity
HF: heart failure
HFpEF: heart failure with preserved ejection fraction
HR: hazard ratio
LAVI: left atrial volume index
LV: left ventricle
LVEDVi: left ventricular end-diastolic volume index
LVEF: left ventricular ejection fraction
LVMI: left ventricular mass index
MI: myocardial infarction
MRA: mineralocorticoid receptor antagonist
NT-proBNP: N-terminal pro–B-type natriuretic peptide
RASi: renin–angiotensin system inhibitor
SGLT2i: sodium-glucose cotransporter-2 inhibitor
sHR: subdistribution hazard ratio
TTE: transthoracic echocardiography

## REFERENCES

1. McDonagh TA, Metra M, Adamo M, Gardner RS, Baumbach A, Böhm M, Burri H, Butler J, Čelutkienė J, Chioncel O, et al. 2021 ESC Guidelines for the diagnosis and treatment of acute and chronic heart failure. Eur Heart J. 2021;42:3599–3726.

2. Heidenreich PA, Bozkurt B, Aguilar D, Allen LA, Byun JJ, Colvin MM, Deswal A, Drazner MH, Dunlay SM, Evers LR, et al. 2022 AHA/ACC/HFSA Guideline for the Management of Heart Failure: a report of the American College of Cardiology/American Heart Association Joint Committee on Clinical Practice Guidelines. J Am Coll Cardiol. 2022;79:e263–e421.

3. Borlaug BA. Evaluation and management of heart failure with preserved ejection fraction. Nat Rev Cardiol. 2020;17:559–573.

4. Shah SJ, Borlaug BA, Kitzman DW, McCulloch AD, Blaxall BC, Agarwal R, Chirinos JA, Collins S, Deo RC, Gladwin MT, et al. Research priorities for heart failure with preserved ejection fraction: National Heart, Lung, and Blood Institute Working Group Summary. Circulation. 2020;141:1001–1026.

5. Pieske B, Tschöpe C, de Boer RA, Fraser AG, Anker SD, Donal E, Edelmann F, Fu M, Guazzi M, Lam CSP, et al. How to diagnose heart failure with preserved ejection fraction: the HFA-PEFF diagnostic algorithm - a consensus recommendation from the Heart Failure Association of the European Society of Cardiology. Eur Heart J. 2019;40:3297–3317.

6. Maurer MS, Burkhoff D, Fried LP, Gottdiener J, King DL, Kitzman DW. Ventricular structure and function in hypertensive participants with heart failure and a normal ejection fraction: the Cardiovascular Health Study. J Am Coll Cardiol. 2007;49:972–981.

7. Lam CS, Roger VL, Rodeheffer RJ, Bursi F, Borlaug BA, Ommen SR, Kass DA, Redfield MM. Cardiac structure and ventricular-vascular function in persons with heart failure and preserved ejection fraction from Olmsted County, Minnesota. Circulation. 2007;115:1982– 1990.

8. Núñez J, Verwerft J, Miñana G, de la Espriella R, Verbrugge FH. Small left ventricular volume in HFpEF: from phenotype to clinical implications. ESC Heart Fail. 2026;13:xvag080.

9. Borlaug BA, Reddy YNV. The role of the pericardium in heart failure: implications for pathophysiology and treatment. JACC Heart Fail. 2019;7:574–585.

10. Felker GM, Ellison DH, Mullens W, Cox ZL, Testani JM. Diuretic therapy for patients with heart failure: JACC state-of-the-art review. J Am Coll Cardiol. 2020;75:1178–1195.

11. Mullens W, Damman K, Harjola VP, Mebazaa A, Brunner-La Rocca HP, Martens P, Testani JM, Tang WHW, Orso F, Rossignol P, et al. The use of diuretics in heart failure with congestion - a position statement from the Heart Failure Association of the European Society of Cardiology. Eur J Heart Fail. 2019;21(2):137–155.

12. Kapelios CJ, Laroche C, Crespo-Leiro MG, Anker SD, Coats AJS, Díaz-Molina B, Filippatos G, Lainscak M, Maggioni AP, McDonagh T, et al. Association between loop diuretic dose changes and outcomes in chronic heart failure: observations from the ESC-EORP Heart Failure Long-Term Registry. Eur J Heart Fail. 2020;22:1424–1437.

13. Rao VN, Pandey A, Zhong L, Ambrosy AP, Fudim M. Loop Diuretic Use and Outcomes in Chronic Stable Heart Failure With Preserved Ejection Fraction. Mayo Clin Proc. 2021;96:503–506.

14. Mullens W, Verbrugge FH, Nijst P, Tang WHW. Renal sodium avidity in heart failure: from pathophysiology to treatment strategies. Eur Heart J. 2017;38:1872–1882.

15. Felker GM, Lee KL, Bull DA, Redfield MM, Stevenson LW, Goldsmith SR, LeWinter MM, Deswal A, Rouleau JL, Ofili EO, et al. Diuretic strategies in patients with acute decompensated heart failure. N Engl J Med. 2011;364:797–805.

16. Lang RM, Badano LP, Mor-Avi V, Afilalo J, Armstrong A, Ernande L, Flachskampf FA, Foster E, Goldstein SA, Kuznetsova T, et al. Recommendations for Cardiac Chamber Quantification by Echocardiography in Adults: An Update from the American Society of Echocardiography and the European Association of Cardiovascular Imaging. J Am Soc Echocardiogr. 2015;28:1–39.

17. Anisman S D, Erickson S B, Morden N E. How to prescribe loop diuretics in oedema BMJ 2019; 364:l359.

18. Popovic D, Alogna A, Omar M, Sorimachi H, Omote K, Reddy YNV, Redfield MM, Burkhoff D, Borlaug BA. Ventricular stiffening and chamber contracture in heart failure with higher ejection fraction. Eur J Heart Fail. 2023;25:657–668.

